# On the Effects of Misclassification in Estimating Efficacy With Application to Recent COVID-19 Vaccine Trials

**DOI:** 10.1101/2020.12.04.20244244

**Authors:** John P. Buonaccorsi

## Abstract

The recent trials for proposed COVID-19 vaccines have garnered a considerable amount of attention and as of this writing extensive vaccination efforts are underway. The first two vaccines approved in the United States are the Moderna and Pfizer vaccines both with estimated efficacy near 95%. One question which has received limited attention, and which we address here, is what affect false positives or false negatives have on the estimated efficacy. Expressions for potential bias due to misclassification of COVID status are developed as are general formulas to adjust for misclassification, allowing for either differential or non-differential misclassification. These results are illustrated with numerical investigations pertinent to the Moderna and Pfizer trials. The general conclusion, fortunately, is that the potential misclassification of COVID status almost always would lead to underestimation of the efficacy and that correcting for false positives or negatives will typically lead to even higher estimated efficacy.

## 1 Introduction

There has been a flurry of recent results coming from large trials of potential COVID-19 vaccines throughout the world. The main object of this note is to explore the potential effects of misclassification of COVID-19 status on the estimated efficacy of the vaccines and assess correction for hypothesized probabilities of false positives and false negatives. This will be done using a combination of analytical results and numerical explorations. Some of the results here have similarities with those in De Smedt et al. [3] who examined the same question in assessing vaccine effectiveness (which as they point out is not the same as efficacy) and connections to the work of Lachenbruch [4], pointed to in more detail later.

In addition to developing some general methodology, we will also illustrate the consequences with numerical investigations specific to the Moderna and Pfizer vaccine trials, both very large studies with volunteers randomized in roughly equal numbers to the vaccine or a placebo. In both studies the primary endpoint was not just whether an individual tested positive but rather if they tested positive and experienced symptoms. We will refer to this combination as “positive” for convenience but in other clinical trials positive may refer to simply being diagnosed positive through testing without symptoms being taken into consideration. Table 1 shows the data for these trials, based on interim analyses used in the FDA approval process, with

**Table 1:** Results from two COVID-19 vaccine trials

| Study | $N_v$ | $N_p$ | $I_v$ | $I_p$ | $R_v$ | $R_p$ | $\widehat{Eff}$ |
| --- | --- | --- | --- | --- | --- | --- | --- |
| Moderna | 13883 | 13934 | 5 | 90 | .00035 | .0065 | 94.44% |
| Pfizer | 17411 | 17511 | 8 | 162 | .00046 | .0093 | 95.04% |

*N*_*v*_ = number of subjects in the vaccinated group

*N*_*p*_ = number of subjects in the placebo group

*I*_*v*_ = number of subjects positive in the vaccinated group

*I*_*P*_ = number of subjects positive in the placebo group

*R*_*v*_ = *I*_*v*_*/N*_*v*_ = positive rate in the vaccinated group

*R*_*p*_ = *I*_*p*_*/N*_*P*_ = positive rate in placebo group

The efficacy of the vaccine is estimated using

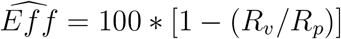

Note that these are interim analyses evaluating efficacy at a fixed point in time after the administration of the vaccine or placebo. We will not discuss methods associated with the time of disease onset (which use proportional hazards models) here. From the statistical perspective the goal is to estimate the true efficacy of the vaccine, defined as

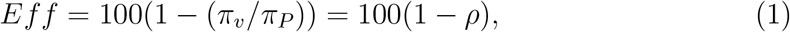

where *ρ* = *π*_*v*_*/π*_*p*_,

*π*_*v*_ = probability a random person from the population is “positive” if vaccinated, and

*π*_*p*_ = probability a random person from the population is “positive” if given the placebo.

We also note that there is no assumption here that the probability a vaccinated person being positive is the same across people, which is not realistic. Each person has a different exposure leading to a different probability and even two people with the exact same exposure could have different probability of being positive. If you vaccinated everyone then the probability a random person selected from the population is positive is *π*_*v*_ which is the average of the individual probabilities over the population. When people are randomly selected and then randomized to the placebo or vaccinated the probability of a vaccinated person being positive (which is over both random selection and assignment) is *π*_*v*_. And given the large samples involved the resulting number of positives, *I*_*v*_, can be treated as Binomial with sample size *N*_*v*_ and probability *π*_*v*_. Similar comments apply for the placebo group.

Assuming, to start, that whether an individual is positive or not can be determined without error, then *R*_*v*_ is unbiased for *π*_*v*_ and *R*_*p*_ is unbiased for *π*_*p*_. Since we are dealing with a ratio however this does not necessarily mean that *R*_*v*_*/R*_*p*_ is unbiased for *ρ* = *π*_*v*_*/π*_*p*_; see the Appendix. For the two trials used for illustration, and for other similarly large trials, any bias from the use of a ratio is extremely small.

A standard error (SE) and confidence interval (CI) for *ρ*, and subsequently for the efficacy, which were absent in the popular reporting of the trial results, can be obtained using methods for estimating a ratio; see [1] and references therein. The resulting standard errors are 2.6% and 1.8% for the Moderna and Pfizer trials respectively with associated confidence intervals of [89.4, 99.4] and [91.3, 98.5], respectively. The confidence intervals are based on the generally more dependable Fieller’s method but these are almost identical to those from the so-called delta method, which use the estimated efficacy ±1.96*SE*. One could also take a Bayesian approach to the problem, which for example leads to a credible region of [90.3, 97.6] as reported for the Pfizer trial, very close to the Fieller interval. To repeat, all of these results are under the assumption that there are no errors in determining whether an individual is positive or not, as defined earlier.

## 2 Bias due to misclassification

There is always the possibility that the assessed status of an individual is incorrect. If the outcome is presence of absence of an infection this arises from errors in the diagnostic procedure; see Lachenbruch [4] for some examples in the diagnosing of Lyme disease. In the two trials here it can arise from misreporting of the presence of absence of symptoms and/or in the diagnosis of presence or absence of COVID from a test. For diagnostic errors it is false negatives that are of the greatest concern with COVID-19.

The natural question is what effect potential misdiagnoses of the primary end-point may have on the estimated efficacy. This is a topic that was certainly absent from the initial coverage of the COVID-19 trials. There is an extensive literature on the potential effects of diagnostic error on the estimation of prevalence, or more generally a proportion, and how to correct for it. In what follows, we draw on the summary in Chapter 2 of Buonaccorsi [2], where numerous additional references can be found.

### 2.1 Non-differential misclassification

To start, assume the sensitivity and specificity are the same in the vaccinated and the placebo group (what is called non-differential misclassification), where with

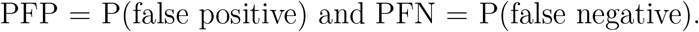

the sensitivity is 1 - PFN and the specificity is 1 - PFP.

The probability of observing a positive result is:

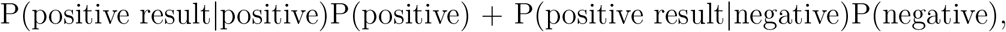

where | denotes “given”. For the vaccinated group this leads to

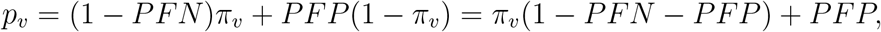

while for the placebo group

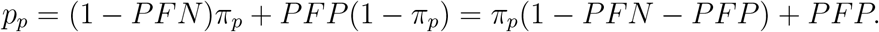

Note that *p* will equal the true *π* if PFN and PFP are both 0 (equivalently, sensitivity and specificity equal 1).

The observed positive rates, *R*_*v*_ and *R*_*p*_ are estimating *p*_*v*_ and *p*_*p*_ respectively, rather than the corresponding *π*_*v*_ and *π*_*p*_ and the observed sample ratio *R*_*v*_*/R*_*p*_ is estimating essentially

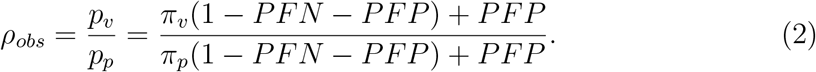

Once again, given that we are dealing with a ratio, this is an approximate/limiting result, not an exact expected value. However, biases based on it are very accurate given large sample sizes, as in the Moderna and Pfizer trials. In the later numerical illustrations this was verified by simulation (results not shown here) for these trials. Note that if one had small sample sizes some modifications will be needed using approximations for expected values of ratios and for also dealing with the fact that there will be a higher probability of a 0 in the denominator, in which case the ratio is not defined. We will not pursue this here.

[Note: Technically, we can only talk about the expected value of the ratio and the subsequent bias in the estimated efficacy if we discard cases where *I*_*p*_ is 0 since the ratio is then undefined. This is not an issue in large trial where the probability that *Ip* = 0 is negiligble. For the Moderna and Pfizer trials this is estimated to be 0 to 40 or 71 decimal places, respecively.]

Defining *c* = 1 − *PFP* − *PFN* for convenience, the bias in the ratio *R*_*v*_*/R*_*p*_ as an estimator of *ρ* = *π*_*v*_*/π*_*p*_ is approximately

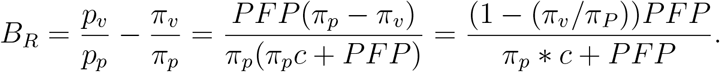

The approximate bias in the estimated efficacy is

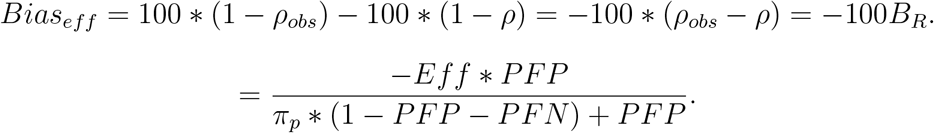

Two main conclusions emerge:

- If the probability of a false positive is 0 then this approximate bias is 0, *regardless of the probability of a false negative*.
- The efficacy will almost always be underestimated. This in contrast to the effect of misclassification in estimating a single proportion where in general the bias may go in either direction depending on the value being estimated and the misclassification probabilities. The only case where the efficacy is underestimated using this result is if the denominator is negative. This cannot happen if 1-PFP - PFN is greater than 0 which would certainly be true for most any sensible assessment procedure.

To explore the issue further, we evaluated the bias numerically over a number of different settings. Based on the results from the two trials we fixed the positive rate in the placebo group, *π*_*p*_, to be .006 to start. We then varied the efficacy over a variety of values between 50% and 95% (which in turn determines the positive rate in the vaccinated group, *π*_*v*_) and used a grid of with the probability of a false positive (PFP) ranging from 0 to .01 and the probability of a false negative (PFN) ranging from 0 to .1 For each combination we evaluated *Eff*_*obs*_ = 100(1 *−* (*p*_*v*_*/p*_*p*_)), which is what is being estimated (approximately) using the observed rates (see earlier results). The process was then repeated by doubling *π*_*p*_ to .012 to reflect higher incidence rates, which would occur over longer observation times, but with the same efficacy.

Figures 1 - 3 display results corresponding to an efficacy of 95%, 70% and 50%, respectively. In each case we plot *Eff*_*obs*_ versus the probability of a false positive (PFP) where the three lines correspond to PFN = .025, .05 and .10. The black lines are for the original setting with *π*_*p*_ = .006 and the red lines for the cases with *π*_*p*_ = .012. The true efficacy is indicated by the solid horizontal line; e.g., at 95 when efficacy = 95%.

**Figure 1:**
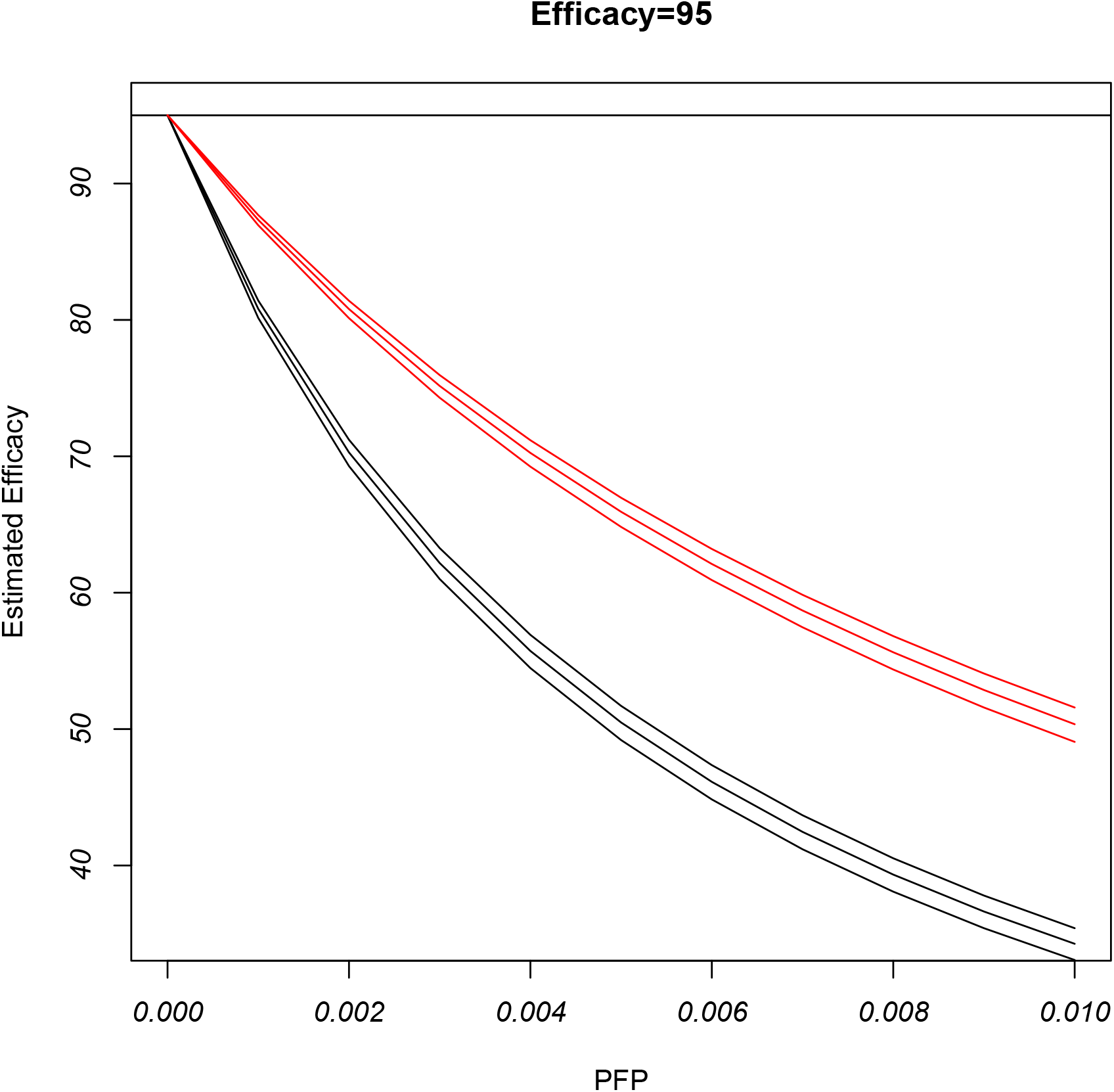
Illustration of bias due to misclassification at efficacy = 95% (solid horizontal line). Plot of what is estimated (on average) using the observed rates as a function of PFP. Black lines are for different PFN (.025, .05 and .10) based on *π*_*p*_ = .006 with *π*_*v*_ = .0003. Red lines are based on *π*_*p*_ = .012 with *π*_*v*_ = .0006.

**Figure 2:**
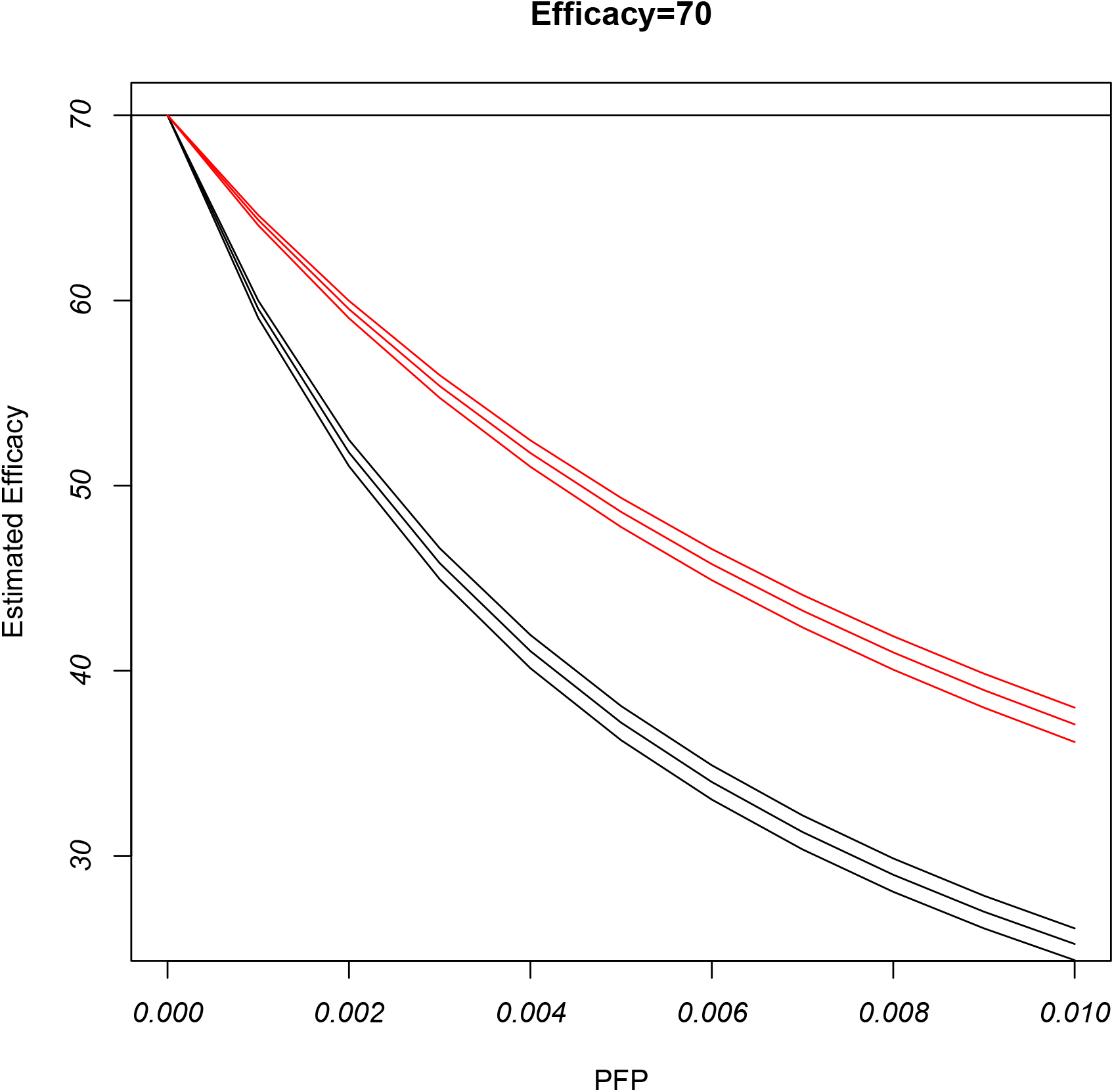
Illustration of bias due to misclassification at efficacy = 70% (solid horizontal line). Plot of what is estimated (on average) using the observed rates as a function of PFP. Black lines are for different PFN (.025, .05 and .10) based on *π*_*p*_ = .006 with *π*_*v*_ = .0018. Red lines are based on *π*_*p*_ = .012 with *π*_*v*_ = .0036.

**Figure 3:**
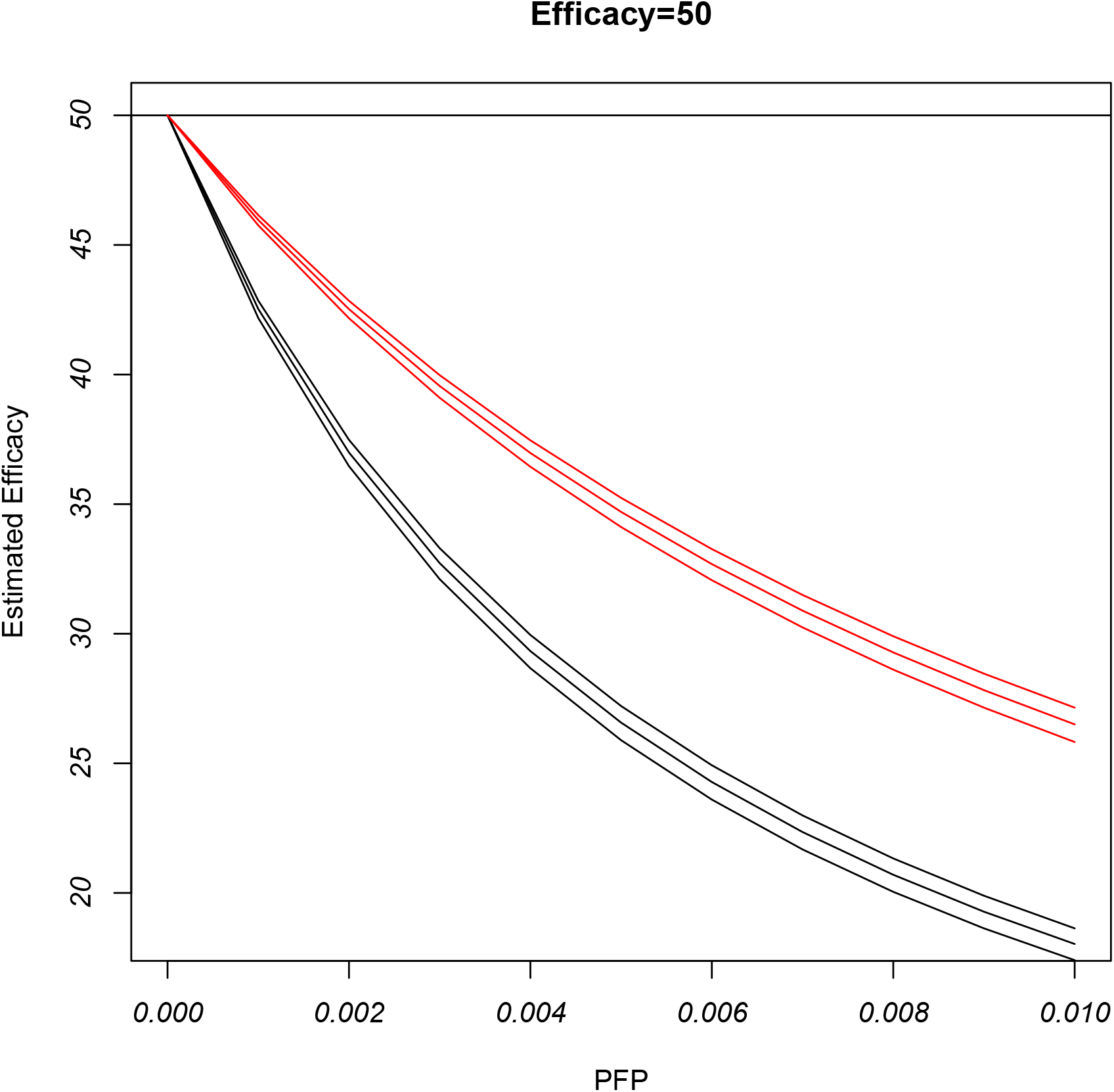
Illustration of bias due to misclassification at efficacy = 50% (solid horizontal line). Plot of what is estimated (on average) using the observed rates as a function of PFP. Black lines are for different PFN (.025, .05 and .10) based on *π*_*p*_ = .006 with *π*_*v*_ = .003. Red lines are based on *π*_*p*_ = .012 with *π*_*v*_ = .006.

As noted earlier the analytical results being used are approximate but simulations using sample sizes of 20000 in each group were in close agreement with those from the approximate expression (results not shown).

A few conclusions emerge immediately from these figures:

1. The efficacy can quickly be grossly underestimated as PFP grows (sensitivity goes down). The rapidity which with this happens is more marked at the smaller rates (*π*_*p*_ = .006 with a corresponding *π*_*v*_ depending on the efficacy.) Frequently in practice, only the smallest PFPs will matter.
2. For a given PFP the bias is rather insensitive to the probability of a false negative (or equivalently to the sensitivity).
3. The bias is greatest at smaller incidence rates (black lines).

Note that the potential biases here are greatest at the rates close to those associated with the Moderna and Pfizer trials.

### 2.2 Adjusting for misclassification

We can also look at adjusting for misclassification and exploring directly what an adjusted estimate of efficacy would look like for the Moderna and Pfizer trials for hypothetical misclassification probabilities.

For given PFP and PFN, corrected estimates of the positive rates, (see for example equation (2.3) in Buonaccorsi [2] or similar equations in Lachenbruch [4]) are given by:

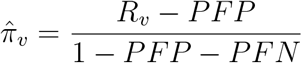

and

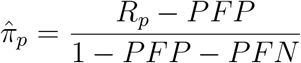

for the vaccinated and placebo groups respectively.

From these a corrected estimate of the efficacy, denoted 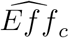 is given by

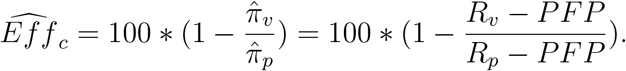

This can also be expressed as

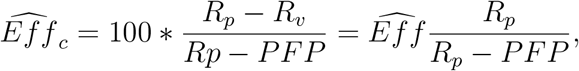

where, recall, 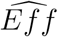 is the original estimate of efficacy based on the observed rates.

- *Although the false negative rate is needed to get a corrected estimate of disease prevalence in each group, it does not enter into getting a corrected estimate of efficacy!* This is similar to what was found in De Smedt et al. [3] for estimating effectiveness. This only applies with non-differential misclassification.
- Note that if *PFP > R*_*v*_ then the corrected estimate of *π*_*v*_ is negative and similarly the estimate of *π*_*p*_ is negative if *PFP > R*_*p*_. Obviously, the underlying probabilities we are trying to estimate cannot be negative. So, with very small estimates of positive rates (as is the case in both the trials being used for illustration) obtaining a corrected estimate will be problematic if the PFP becomes too large. This would occur (i.e., both estimates would be negative) if PFP is greater .006 in the Moderna trial or greater than .008 in the Pfizer trial.

We applied the correction described to the original data for varying levels of PFP (chosen to keep the corrected estimated rates positive) for each of the trials with results as shown in the black lines in Figures 4 and 5. The red lines in those figures come from doubling the positive rates in the original data to see to what extent the very small rates in the original data influenced the conclusions. Recall the probability of a false negative does not enter into getting a corrected estimate of efficacy under non-differential misclassification.

**Figure 4:**
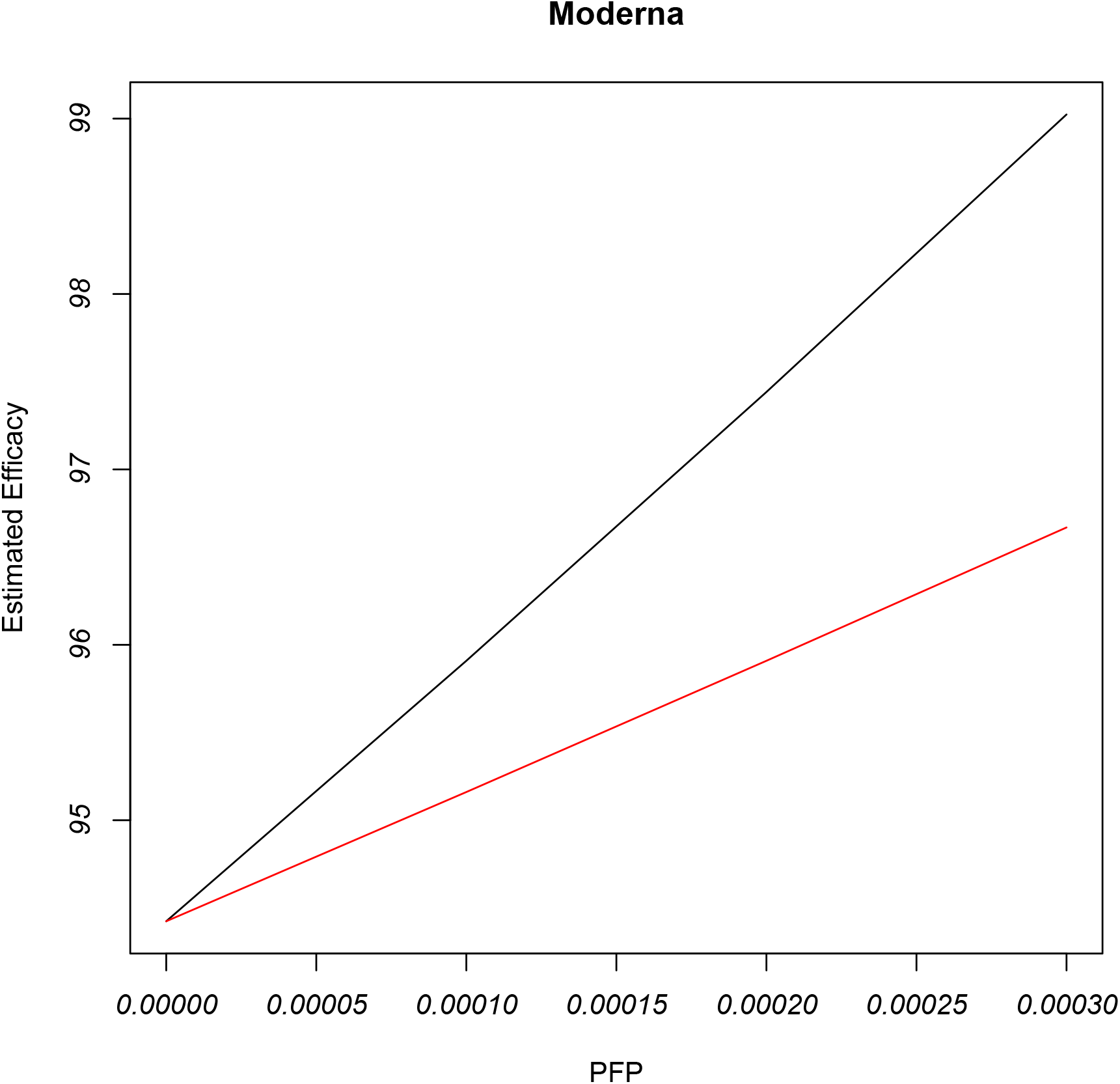
Illustration of correction of estimated efficacy with the Moderna data as a function of possible values of PFP. Black line uses the original data while red line is with doubled positive rates. Original estimated efficacy equals 94.4% in each case

**Figure 5:**
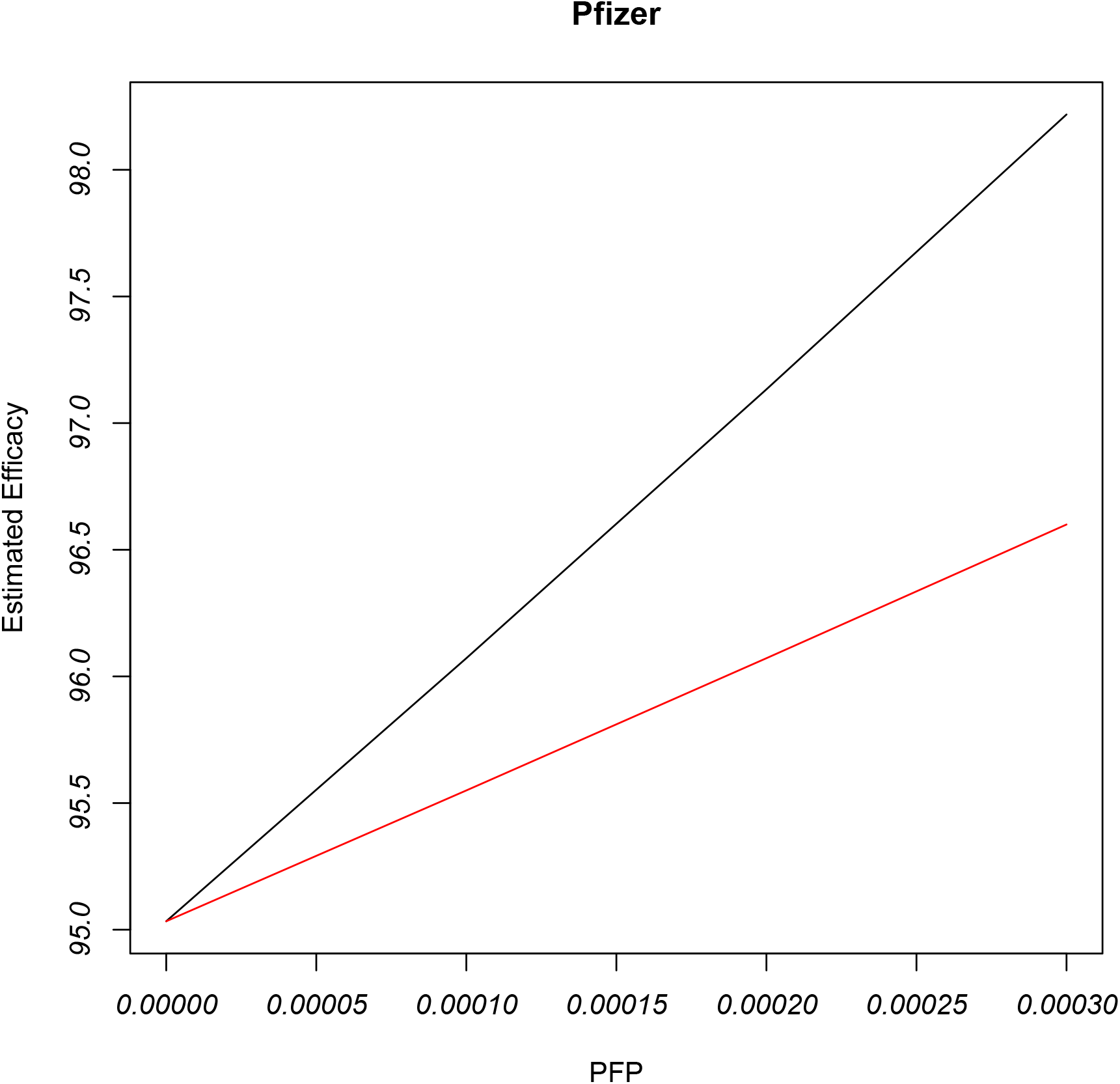
Illustration of correction of estimated efficacy with the Pfizer data as a function of possible values of PFP. Black line uses the original data while red line is with doubled positive rates. Original estimated efficacy equals 95.04% in each case

The story is pretty clear here and was expected based on the earlier theoretical bias discussion. Correcting for misclassification due to non-zero probability of a false positive always leads to a larger estimated efficacy. For these two trials even a modest PFP leads to estimates approaching 100%.

### 2.3 Differential Misclassification

#### 2.3.1 Biases

Differential misclassification occurs if the probabilities of false positives and false negatives differ in the placebo and vaccinated groups. In this case the analytical assessment of bias induced by misclassification becomes more complicated. We now denote the probabilities involved by:

*PFP*_*v*_ = P(false positive) in vaccinated group

*PFN*_*v*_ = P(false negative) in vaccinated group

*PFP*_*p*_ = P(false positive) in placebo group, and

*PFN*_*p*_ = P(false negative) in placebo group.

The estimator based on the observed positive rates, 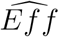, is now is estimating

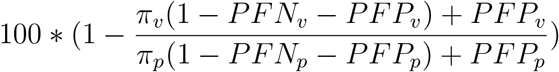

rather than the true efficacy. This leads to a bias of

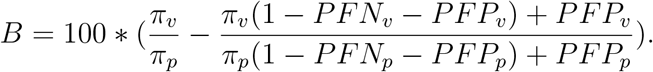

Unlike the case with non-differential misclassification there is no simple expression leading to easy insight into the nature of the bias. However it is easy to explore numerically. Similar to what was done earlier under non-differential misclassification, the true positive rate in the placebo group was set to .006 with efficacies of 95% (*π*_*v*_ = .0003),70% (*π*_*v*_ = .0018) and 50% (*π*_*v*_ = .006). We looked at many combinations resulting from varying *PFP*_*v*_ and *PFP*_*p*_ from 0 to .01 and *PFN*_*v*_ and *PFN*_*p*_ from 0 to .10. Figures 6 - 8 show the nature of the bias as a function of *PFP*_*v*_ for specified values of *PFP*_*p*_ not equal to *PFP*_*v*_. The plots show what is being estimated using the observed rates compared to the true efficacy (indicated by the solid black horizontal line) where at each value of *PFP*_*v*_ the multiple values correspond to all the combinations of the two probabilities of false negatives, some of which are equal, others which are not.

**Figure 6:**
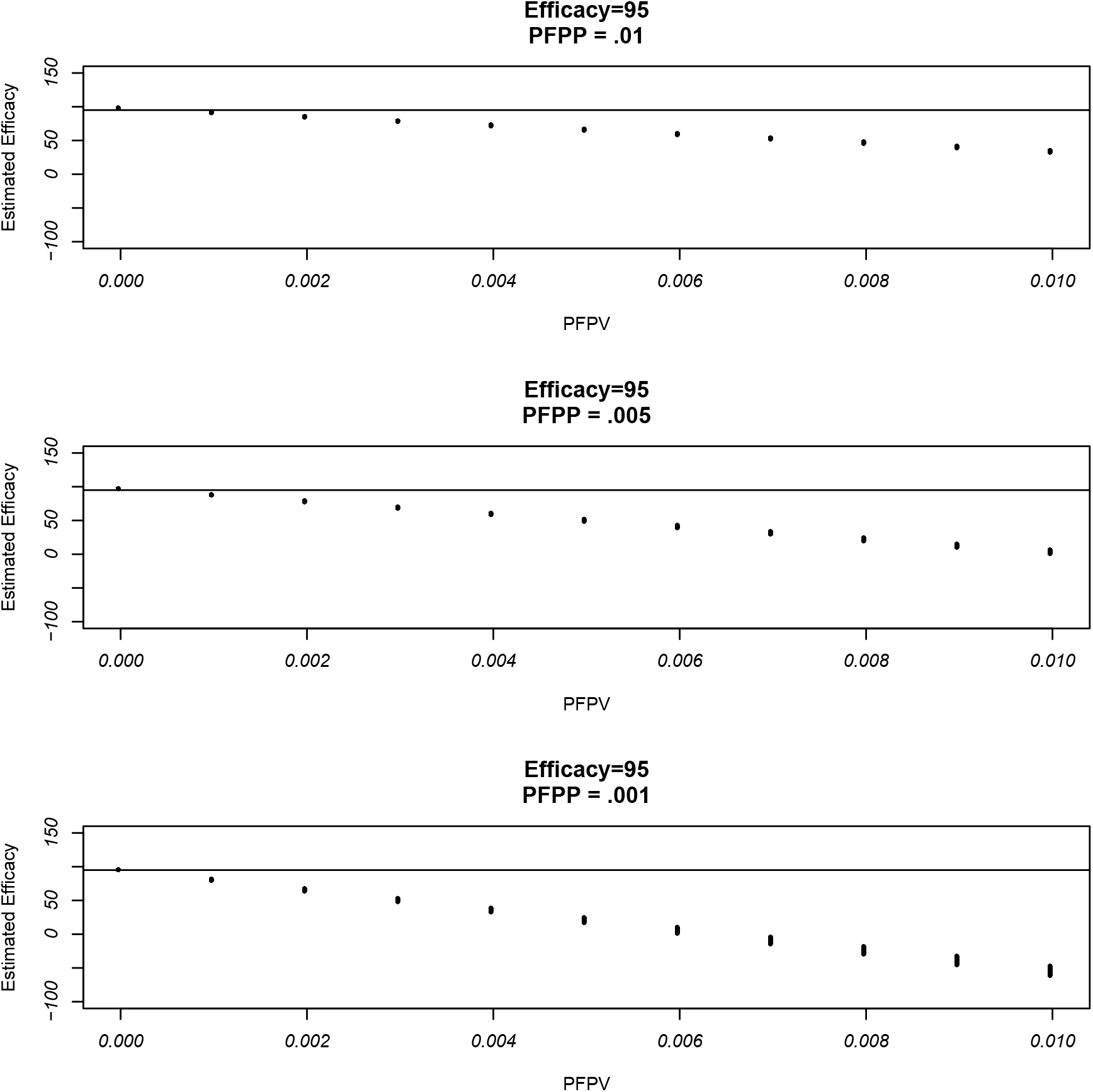
Illustration of bias due to differential misclassification at efficacy = 95% (solid horizontal line). PFPV = probability of false positive in vaccinated group. PFPP = probability of false positive in placebo group. Plot of what is estimated (on average) using the observed rates as a function of PFPV with different points from combinations of probabilities of false negatives over both groups. Based on *π*_*p*_ = .006 and *π*_*v*_ = .0003.

**Figure 7:**
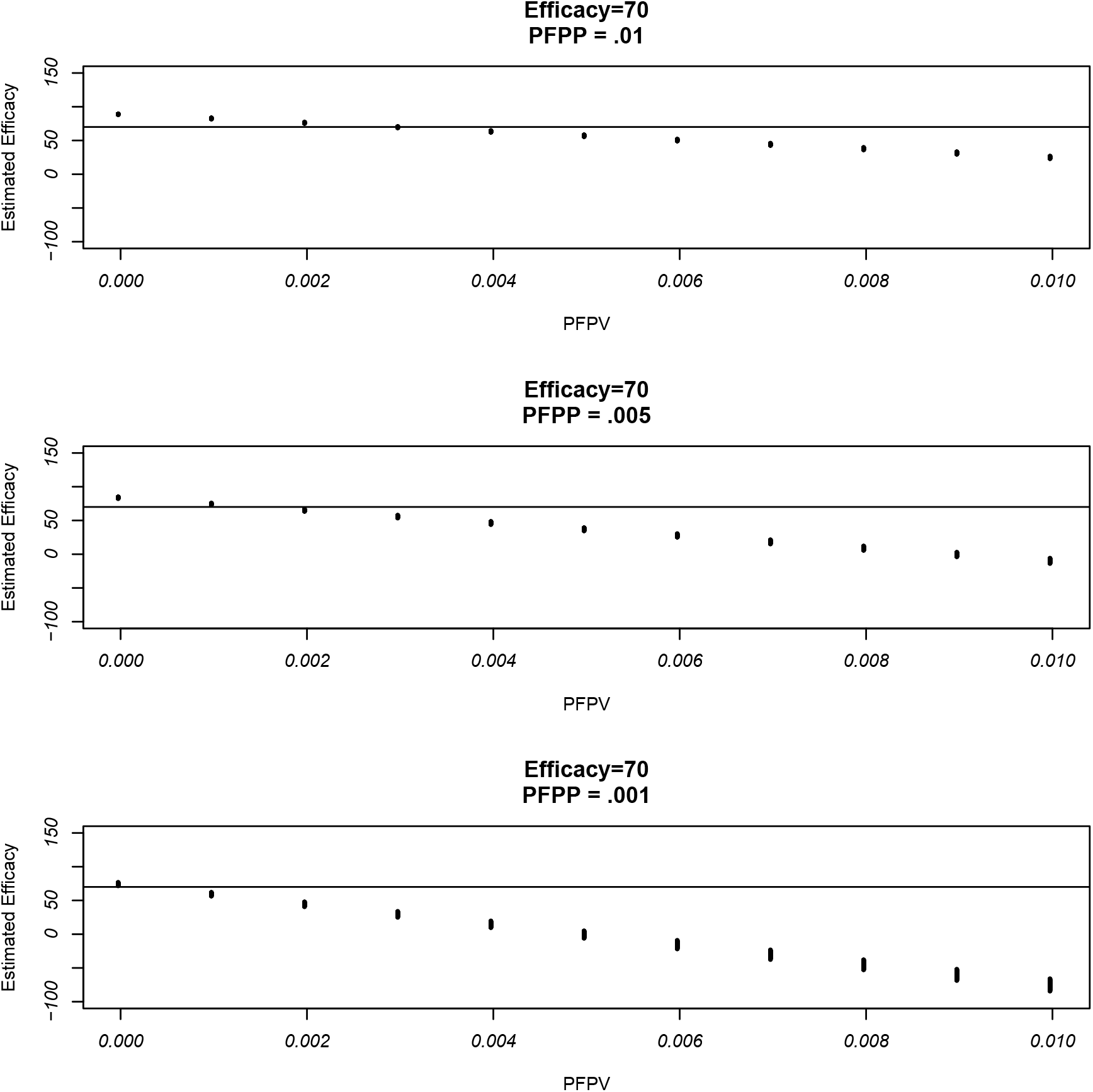
Illustration of bias due to differential misclassification at efficacy = 70% (solid horizontal line). PFPV = probability of false positive in vaccinated group. PFPP = probability of false positive in placebo group. Plot of what is estimated (on average) using the observed rates as a function of PFPV with different points from combinations of probabilities of false negatives over both groups. Based on *π*_*p*_ = .006 and *π*_*v*_ = .0018.

**Figure 8:**
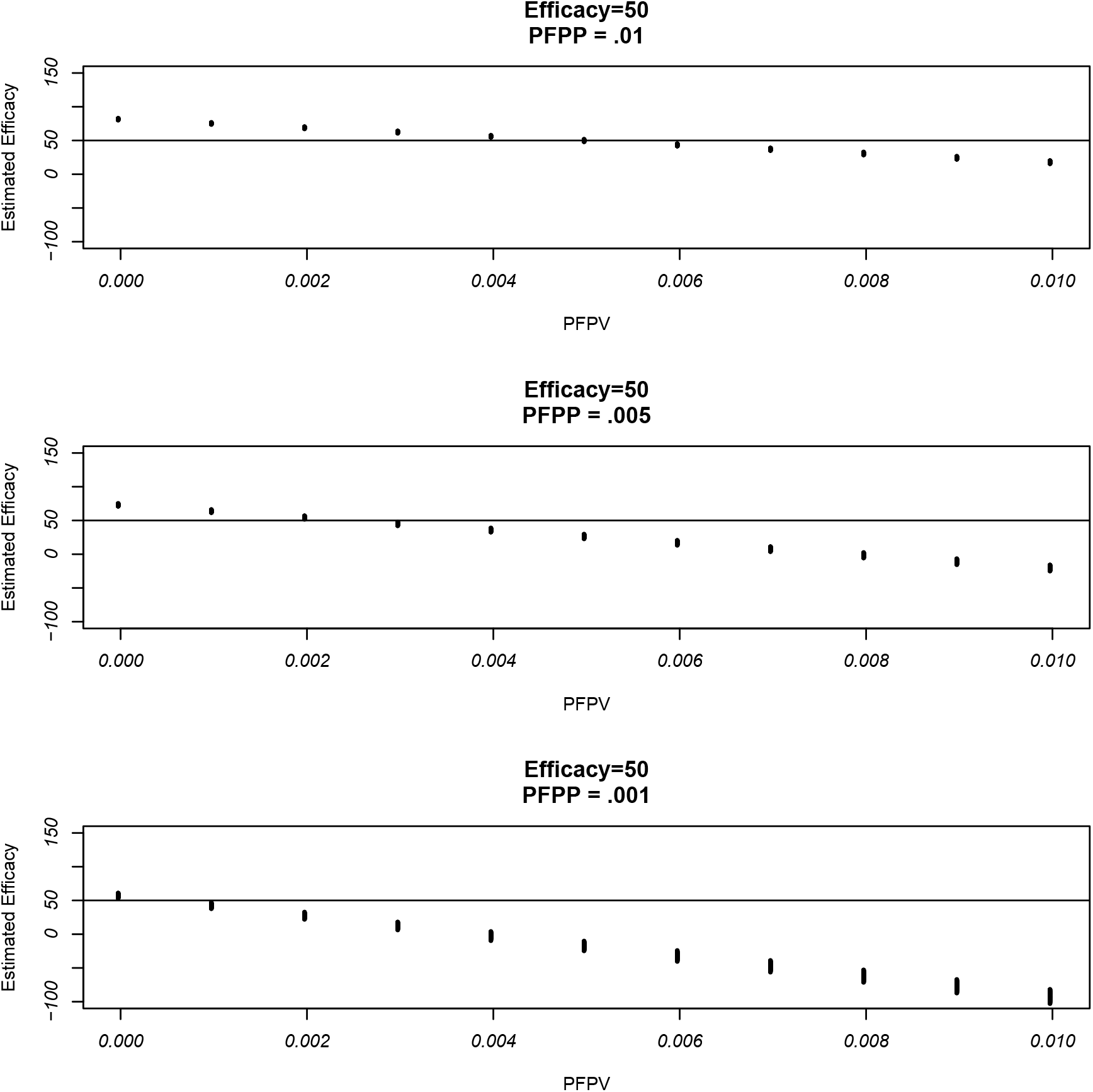
Illustration of bias due to differential misclassification at efficacy = 50% (solid horizontal line). PFPV = probability of false positive in vaccinated group. PFPP = probability of false positive in placebo group. Plot of what is estimated (on average) using the observed rates as a function of PFPV with different points from combinations of probabilities of false negatives over both groups. Based on *π*_*p*_ = .006 with *π*_*v*_ = .003.

- The nature of the bias is determined primarily by the probabilities of false positives involved.
- The nature of the bias is not very sensitive to the probabilities of false negatives, although, interestingly, the spread of biases of the different probabilities of false negatives is greater the smaller the probability of a false positive is in the placebo group.
- The broad message is that as with non-differential misclassification the concern is primarily with underestimation of the efficacy. At the higher efficacies (95% and 70%) the efficacy is almost always underestimated with the exception being a few unlikely settings where the PFP in the vaccinated group is 0 or very small and probability of a false positive in the placebo groups becomes larger than it. But even then the overestimation is slight.

At an efficacy of 50% there can be more in the way of potential overestimate in certain situations, although this scenario is not relevant for the Pfizer and Moderna trials. As above, this happens when the probability of a false positive in the is vaccinated group is low and the probability of a false positive in the placebo group is higher. Then the overestimation could be concerning although this is probably an unlikely scenario and we note that at the smaller PFP of .001 in the placebo group, the efficacy will be underestimated.

#### 2.3.2 Correcting for misclassification

The general strategy of correcting for misclassification using information on the false positive and false negative rates is similar to the non-differential cases. The corrected estimates of the positive rates in the two groups are

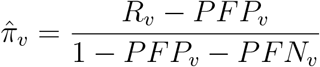

for the vaccinated group and

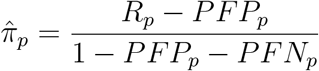

for the placebo group.

From these the corrected estimate of the efficacy, denoted 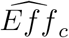 is given by

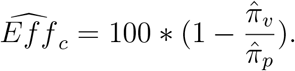

Similar to treating bias, there are no simple expressions showing the nature of the correction and unlike the case of non-differential misclassification we do need information about both the false positive and false negative rates, for each group. This was also noted by De Smedt et al. [3] for estimating effectiveness.

Figs 9 and 10 show the effects of correcting for misclassification using the Moderna and Pfizer data using all combination of probabilities of false positives ranging from 0 to .0003 in each group and probabilities of false negatives in the two groups ranging from 0 to .01. As with non-differential misclassification, we see that the correction almost always moves the original estimated efficacy closer to 100%. The only exception to this were very small drops in the estimated efficacy in the unrealistic scenarios where the PFP in the vaccinated group was 0 but was positive in the placebo group.

**Figure 9:**
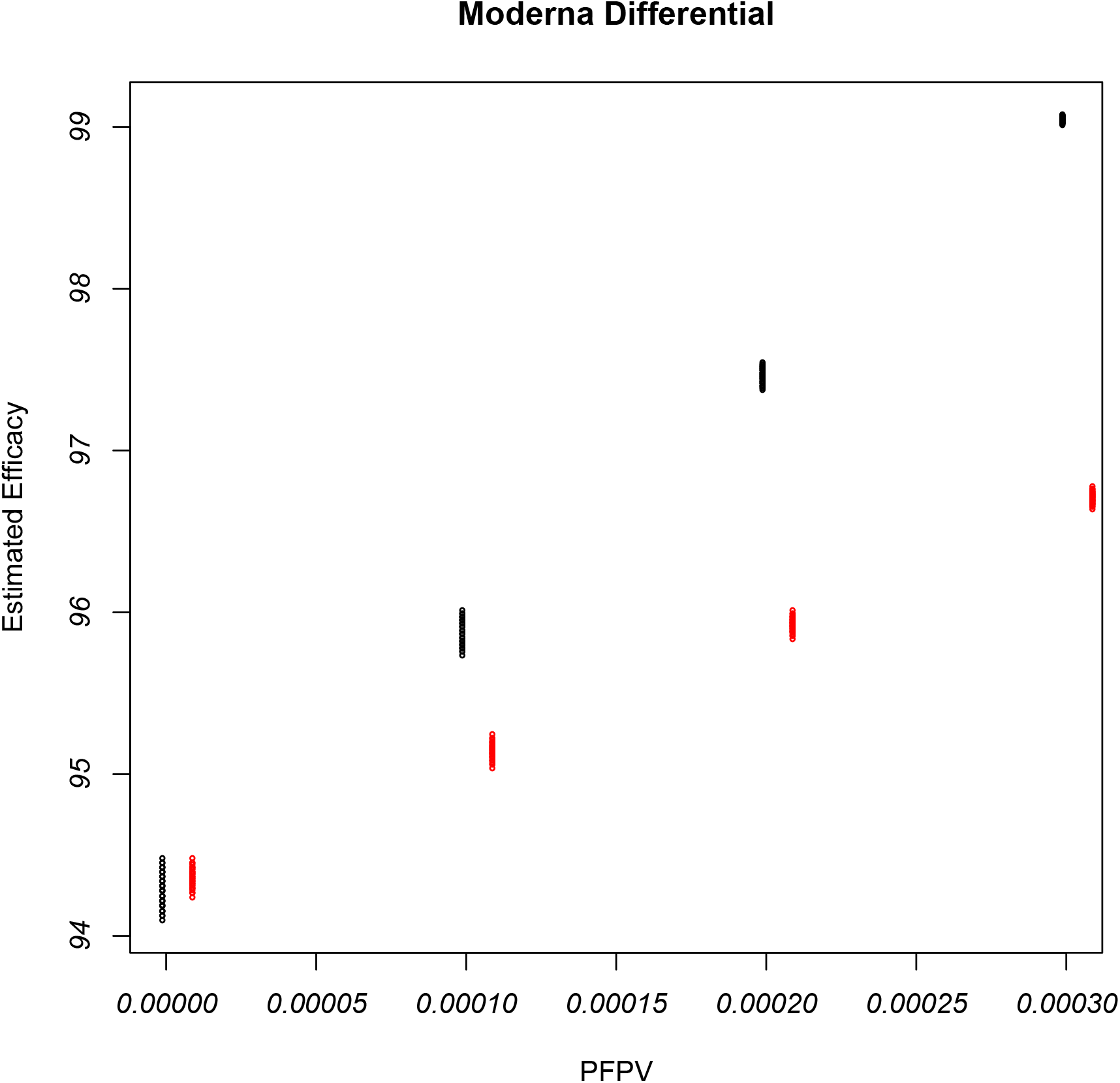
Illustration of corrected estimated efficacy with the Moderna data as a function of possible values of PFPV = probability of false positive in vaccinated group with differential misclassification. Black points are for the original data and range over all values of PFP in placebo groups and PFNs in the two groups. Red points (offset slightly for plotting purposes) use double the original positive rates. Original estimated efficacy equals 94.4% in each case.

**Figure 10:**
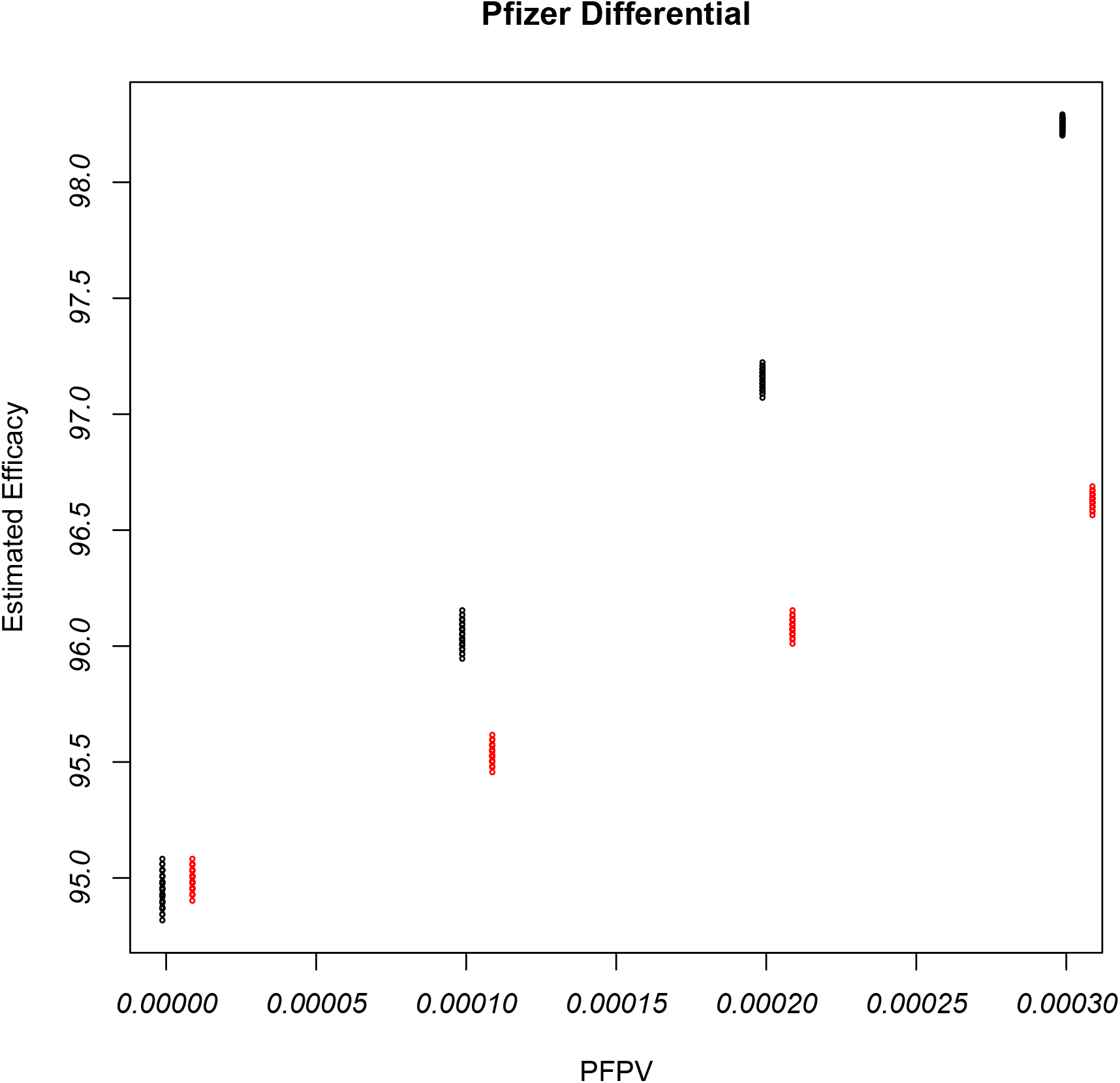
Illustration of corrected estimated efficacy with the Prizer data as a function of possible values of PFPV = probability of false positive in the vaccinated group with differential misclassification. Black points are for the original data and range over all values of PFP in placebo groups and PFNs in the two groups. Red points (offset slightly for plotting purposes) use double the original positive rates. Original estimated efficacy equals 95.04% in each case.

## 3 Discussion

The main goal here was impact of potential misclassification of the primary endpoint in estimating efficacy and how one can correct the estimated efficacy using information about misclassification rates. The general conclusion is that in the majority of settings (including that of the Moderna and Pfizer trials) the misclassification would, on average, lead to underestimation of the efficacy and correcting for misclassification would push the estimate efficacy higher towards 100%.

A fuller treatment of the problem would also address the issue of how to get standard errors and confidence intervals for the estimated efficacy using either known or estimated probabilities of false positives and false negatives. This can be handled in a relatively straightforward manner using inferences for ratios. In the case of using estimated misclassification rates, the particulars of the analysis will depend on whether the validation is internal or external. This is tangential to the main focus here and will not be addressed in detail here.

## Data Availability

Data taken off web and public press reporting COVID-19 vaccine trials by Moderna and Pfizer

## Appendix

It is well known that the expected value of a ratio is not exactly the ratio of the expected value. (Recall, that as noted in the text that it only makes sense to talk about the expected value of the ratio if the probability that the denomiator is 0, is essentially 0). So, in our setting with no misclassification, the expected value of *R*_*v*_*/R*_*p*_ is not exactly *π*_*v*_*/π*_*v*_. But, using an approximation (see for example page 181 of Mood, Graybill and Boes [4]) in this situation, where *R*_*p*_ and *R*_*v*_ are uncorrelated, the bias in the ratio is approximately 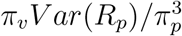. Using the data this can be estimated by 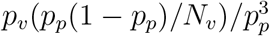, which will be small for large *N*_*v*_. The estimated bias in the efficacy is −100*(bias in the ratio). This is estimated to be -.062 and -.03 for the Moderna and Pfizer trials respectively, which for all practical purposes is 0 compared to the estimated efficacies near 95. When, misclassification is present one can use a similar argument to show that the difference between *E*(*R*_*v*_*/R*_*p*_) and *p*_*v*_*/p*_*v*_ is negligible.

## Notes

### Competing Interest Statement

The authors have declared no competing interest.

### Funding Statement

No external funding received.

### Author Declarations

No IRB oversight involved

### Summary of Updates

- Clarified language throughout the manuscript to generally refer to positive outcome; which in the Moderna and Pfizer trials means symptoms plus a positive diagnosis but in other cases may refer simply to a positive diagnosis through some testing mechanism. - Correction of a few typos and misspellings. - Clarification in captions to figures 1,2,3,6,7 and 8 that y axis is giving what is estimated “on average”. - Added Lachenbruch reference

